# Inequalities in Compliance with the Two-Dose Measles Vaccination in Peruvian Children Aged 12 to 59 Months in 2023

**DOI:** 10.1101/2024.06.03.24308410

**Authors:** Claudio Intimayta-Escalante, Gustavo Tapia-Sequeiros, Luis E. Cueva-Cañola

**Author notes:** **Corresponsal Author: Claudio Intimayta-Escalante, E-mail:**.

## Abstract

**Introduction:** Inadequate measles vaccination coverage leads to outbreaks that affect the pediatric population in Peru. The aim of this study was to evaluate sociodemographic inequalities related to the coverage and compliance in the two-dose measles vaccination schedule among Peruvian children in 2023.

**Methods:** An analytical cross-sectional study was conducted using data from a national Peruvian survey. Mothers aged 15 to 49 with children aged 12 to 59 months were included. Sociodemographic and health characteristics of the mother and child were considered sources of inequality. Prevalence ratios (PR) were calculated to measure the characteristics associated with adherence to the measles vaccination schedule. Inequality analysis was assessed using the Erreygers Concentration Index (ECI).

**Results:** A total of 8,148 households were evaluated. Vaccination coverage was identified as 85.9% (95%CI: 94.7 – 87.1) for the first dose and 53.9% (95%CI: 52.1 – 55.7) for the second dose. In the inequality analysis, mothers with no education (CI: -0.57), aged 15 to 17 years (CI: -0.38), residing in the jungle (CI: -0.41) or highlands (CI: -0.26), and without health insurance (CI: -0.01) showed greater inequality in adherence to the vaccination schedule. Additionally, adherence to growth and development check-ups in infants was a source of inequality (CI: -0.09).

**Conclussion:** Measles vaccination coverage did not reach the minimum values required to prevent outbreaks in Peru. The factors contributing to inequality in vaccination coverage included education level, age, region of residence, area of residence, health insurance status, and adherence to growth and development check-ups.

## INTRODUCTION

Immunizations are one of the most effective measures for preventing diseases, as they prevent between 2 and 3 million deaths annually worldwide (1-3). Measles is highly contagious among the pediatric population and presents with symptoms such as diarrhea, ear infections, blindness, pneumonia, encephalitis, and, in some cases, can lead to death (4). In many countries, including the United States and the United Kingdom, the measles, mumps, and rubella (MMR) vaccine is distributed free of charge (5). The first dose is typically administered at 12 months of age, with a booster dose given at 6 years of age, following the recommended vaccination schedule.

In 2019, the World Health Organization (WHO) estimated 869,770 cases and 207,500 deaths due to measles (7). The global immunization rate declined by 5% from 2019 to 2021, possibly due to factors such as vaccine hesitancy, disruptions in healthcare services, or logistical challenges. This situation, exacerbated by the COVID-19 pandemic, primarily affected low- and middle-income countries (8). Only 7 out of 40 countries attained a minimum coverage of 95% to prevent measles outbreaks in 2024, highlighting the persistent risk of widespread outbreaks in regions with low vaccination rates.

In Peru, immunization coverage against measles was estimated at 70.2% and 52% for the first and second doses, respectively, in 2017 (11). However, due to reduced immunization coverage during the pandemic, measles cases became more frequent (12, 13). Low vaccination rates can be attributed to public unawareness, such as misinformation spread on social media. In addition, inadequate healthcare supplies, like shortages of vaccines or insufficient training of healthcare personnel, can lead to errors in vaccine administration (14). However, there is no specific evidence regarding how sociodemographic conditions influence inequality in completing the immunization schedule. Therefore, the aim of this study was to evaluate sociodemographic inequalities associated with coverage and adherence to the two-dose measles vaccination schedule among Peruvian children aged 12 to 59 months in 2023.

## METHODS

### Study Design

We conducted an analytical cross-sectional study using data from the 2023 Demographic and Family Health Survey (or ENDES, in Spanish acronym) (15). The National Institute of Statistics and Informatics (or INEI, in Spanish acronym) conducts the ENDES survey every year since 2004 in urban and rural areas throughout Peru’s regions, including the constitutional province of Callao. Mothers aged 15 to 49 years with Peruvian children aged 12 to 59 months who responded to questions about sociodemographic charancteristics and health issues like insurance affiliation, prenatal care visits, and immunization were also evaluated.

### Variables Related to the Mothers

Sociodemographic and health characteristics were evaluated, such as age group (18 to 24, 25 to 34, 35 to 49 years), educational level (no education, primary, secondary, or higher), wealth index (first, second, third, fourth, and last quintile), area of residence (rural and urban), and natural region of residence (Lima, rest of the coast, highlands, and jungle). Additionally, health-related aspects, including insurance affiliation (yes or no) and receipt of tetanus vaccination during pregnancy, were examined. The assessment included identifying if the childbirth took place at a health center (yes or no).

### Variables Related to the Child

Additionally, the evaluation included determining if children received the initial dose of measles vaccination and the booster or second dose of measles vaccination. A child was considered to have completed both doses if the vaccination was recorded on the card, with the date documented, or if the mother reported the immunization. The child’s age in months was assessed in ranges (12 to 23 months, 24 to 35 months, 36 to 47 months, and 48 to 59 months), along with evaluating if they received growth and development check-ups (yes or no).

### Statistical Analysis

The statistical analysis was conducted using STATA version 17.0, incorporating the complex sample design of the ENDES. Categorical variables were described using frequencies and percentages, along with their respective 95% confidence intervals weighted by the design effect. The study used the Rao-Scott chi-square test to analyze differences in the proportion of children who completed both doses of measles vaccination across different categories of the study variables. The analysis aimed to explore the connection between the assessed characteristics of mothers and children and the completion of both measles vaccination doses. We used Poisson regression models with strong variances to find the crude prevalence ratio (cPR) and the adjusted prevalence ratio (aPR), taking into account all the others characteristics.

### Inequality Analysis

The inequality analysis was conducted using concentration curves to assess the compliance of the doses of measles vaccination based on socioeconomic status, ranging from the poorest to the richest. The concentration index (CI) was calculated by evaluating the area above or below the concentration curve. Poverty-related inequality is associated with situations that lean toward accumulation above the curve (16). Additionally, the Erreygers Concentration Index (ECI) was used to ensure a fairer calculation by considering the extremes of the evaluated health service distribution. Also, maps were generated to depict the inequality in immunization coverage among the regions of Peru.

### Ethical Aspects

The study was conducted by analyzing data from ENDES, a national survey administered with informed consent from participants. Additionally, the study did not have information that would allow the identification of participants included in the research.

## RESULTS

Among the 8,148 households included in the study, a coverage of 85.9% (95%CI: 84.7 – 87.1) for the first dose and 53.9% (95%CI: 52.1 – 55.7) for the booster dose of the measles vaccine was identified. Evaluating maternal characteristics revealed that sociodemographic factors (age group, educational level, wealth index, natural region, and area of residence) and health factors during pregnancy (prenatal care, receipt of tetanus vaccine, and institutional childbirth) mediated significant differences in the proportion of vaccinated infants (p*<*0.05). Similarly, infant age and participation in growth and development check-ups were found to determine significant differences in the proportion of vaccinated infants (p*<*0.05).

Additionally, it was identified that 52.1% (95%CI: 50.3 – 54.0) of infants completed both doses of vaccination. Evaluating maternal characteristics revealed that maternal age group, educational level, natural region, and area of residence, as well as receiving prenatal care, tetanus vaccination during pregnancy, and institutional childbirth, mediated significant differences in the proportion of infants vaccinated with the first dose (p*<*0.05). Furthermore, infant age and participation in growth and development check-ups were found to mediate differences in vaccination proportion among these infants (p*<*0.05). However, this was not the case for the proportion of infants vaccinated with the booster dose, among whom only certain maternal characteristics such as age group, area of residence, receiving prenatal care, tetanus vaccination during pregnancy, and institutional childbirth mediated significant differences (p*<*0.05). Similarly, this was observed for infant age in months within this group (Table 1).

**Table 1.** Characteristics of mothers and children according to coverage of the first and second doses of measles vaccine.

| Variables | First Doses (n=6802) |  | P-value** | Second Doses (n=4095) |  | P-value** |
| --- | --- | --- | --- | --- | --- | --- |
|  | Proportion<br>according to<br>characteristics | First dose<br>coverage*<br>(12 to 59 months) |  | Proportion<br>according to<br>characteristics | Second dose<br>coverage*<br>(12 to 59 months) |  |
|  | %* (95%CI) | %* (95%CI) |  | %* (95%CI) | %* (95%CI) |  |
| **Mother's Characteristics** |  |  |  |  |  |  |
| **Age Group** |  |  |  |  |  |  |
| 15 to 17 years old | 0.63 (0.4 a 0.9) | 67.2 (46.2 a 83.0) | <0.001 | 0.4 (0.2 a 0.9) | 27 (13.0 a 47.8) | <0.001 |
| 18 to 29 years old | 37.2 (35.4 a 39.0) | 82.3 (80.2 a 84.3) |  | 36.4 (34.2 a 38.6) | 50.5 (47.7 a 53.2) |  |
| 30 to 49 years old | 62.2 (60.4 a 64.0) | 88.5 (87.1 a 89.7) |  | 63.2 (61.0 a 65.4) | 56.4 (54.2 a 58.6) |  |
| **Educational Level** |  |  |  |  |  |  |
| No Education | 0.8 (0.6 a 1.1) | 79 (65.7 a 88.0) | <0.001 | 0.9 (0.6 a 1.3) | 54.9 (41.1 a 67.9) | 0.605 |
| Elementary | 11.4 (10.4 a 12.5) | 82.2 (78.9 a 85.0) |  | 11.9 (10.6 a 13.3) | 53.5 (49.5 a 57.5) |  |
| High School | 45.6 (43.7 a 47.6) | 84.4 (82.5 a 86.0) |  | 45.7 (43.3 a 48.2) | 52.9 (50.4 a 55.4) |  |
| University | 42.2 (40.3 a 44.2) | 88.9 (87.0 a 90.6) |  | 41.5 (39.1 a 44.0) | 55.1 (52.1 a 58.1) |  |
| **Wealth Index** |  |  |  |  |  |  |
| Q5 (Last Quintile) | 18.5 (17.5 a 19.7) | 81.2 (78.8 a 83.3) | 0.001 | 18.5 (17.1 a 19.9) | 50.5 (47.7 a 53.3) | 0.32 |
| Q4 | 23.1 (21.7 a 24.6) | 85.1 (82.8 a 87.2) |  | 23.2 (21.5 a 25.1) | 53.6 (50.5 a 56.6) |  |
| Q3 | 20.1 (18.6 a 21.7) | 86.4 (83.5 a 88.9) |  | 20.1 (18.2 a 22.2) | 54.2 (50.6 a 57.8) |  |
| Q2 | 20.3 (18.7 a 22.0) | 87.7 (84.5 a 90.3) |  | 20.4 (18.4 a 22.5) | 55.6 (51.3 a 59.7) |  |
| Q1 (First Quintile) | 18.0 (16.3 a 19.8) | 89.7 (86.1 a 92.5) |  | 17.8 (15.9 a 19.9) | 55.9 (50.4 a 61.3) |  |
| **Natural Region** |  |  |  |  |  |  |
| Metropolitan Lima | 36.0 (34.3 a 37.8) | 87.7 (85.0 a 90.1) | 0.008 | 35.4 (33.0 a 37.9) | 54.3 (50.1 a 58.4) | 0.216 |
| Rest of the Coast | 26.1 (24.8 a 27.5) | 86.8 (84.6 a 88.7) |  | 27 (25.2 a 28.9) | 56.2 (53.4 a 59.0) |  |
| Highlands | 23.8 (22.5 a 25.2) | 84.9 (83.0 a 86.6) |  | 23.6 (21.9 a 25.4) | 52.6 (50.1 a 55.1) |  |
| Jungle | 14.0 (13.1 a 15.0) | 81.8 (79.5 a 83.8) |  | 14 (12.7 a 15.3) | 51 (48.1 a 54.0) |  |
| **Area of Residence** |  |  |  |  |  |  |
| Urban | 83.5 (82.52 a 84.40) | 86.7 (85.3 a 88.0) | <0.001 | 83.9 (82.6 a 85.1) | 54.7 (52.6 a 56.8) | 0.01 |
| Rural | 16.5 (15.60 a 17.48) | 82.3 (80.0 a 84.3) |  | 16.1 (14.9 a 17.4) | 50.1 (47.3 a 52.9) |  |
| **Health Insurance** |  |  |  |  |  |  |
| Yes | 93.4 (92.3 a 94.3) | 86.2 (85.0 a 87.4) | 0.037 | 93.6 (92.2 a 94.8) | 54.3 (52.4 a 56.1) | 0.137 |
| No | 6.6 (5.7 a 7.7) | 81.7 (76.6 a 85.8) |  | 6.4 (5.2 a 7.8) | 48.9 (42.0 a 55.8) |  |
| **Antenatal Care Visits** |  |  |  |  |  |  |
| Less than Six | 12.1 (10.8 a 13.5) | 77 (72.5 a 80.9) | <0.001 | 11.4 (9.9 a 13.0) | 45.9 (40.9 a 51.0) | <0.001 |
| Six or More | 87.9 (86.6 a 89.2) | 89.1 (87.9 a 90.3) |  | 88.6 (87.0 a 90.1) | 56.7 (54.8 a 58.7) |  |
| **Received Tetanus Vaccine** |  |  |  |  |  |  |
| No | 19.8 (18.3 a 21.5) | 80.9 (77.6 a 83.8) | <0.001 | 18.3 (16.4 a 20.3) | 47.1 (43.3 a 50.9) | <0.001 |
| Yes | 80.2 (78.5 a 81.7) | 89.1 (87.8 a 90.3) |  | 81.7 (79.7 a 83.6) | 57.4 (55.4 a 59.5) |  |
| **Institutional Labor** |  |  |  |  |  |  |
| No | 3.8 (3.2 a 4.6) | 70.3 (63.2 a 76.5) | <0.001 | 3.2 (2.6 a 3.9) | 36.6 (30.5 a 43.3) | <0.001 |
| Yes | 96.2 (95.4 a 96.8) | 86.7 (85.5 a 87.8) |  | 96.8 (96.1 a 97.4) | 54.7 (52.9 a 56.5) |  |
| **Children's Characteristics** |  |  |  |  |  |  |
| **Age (months)** |  |  |  |  |  |  |
| 12 to 23 | 20.4 (19.0 a 21.8) | 78.7 (75.7 a 81.4) | <0.001 | 12.3 (10.9 a 13.8) | 29.9 (26.8 a 33.2) | <0.001 |
| 24 to 35 | 23.1 (21.6 a 24.7) | 86.8 (84.3 a 89.0) |  | 26.6 (24.7 a 28.7) | 62.7 (59.1 a 66.1) |  |
| 36 to 47 | 27.9 (26.2 a 29.6) | 88.8 (86.7 a 90.6) |  | 30.3 (28.1 a 32.6) | 60.6 (57.4 a 63.7) |  |
| 48 to 59 | 28.6 (27.0 a 30.3) | 88.2 (86.2 a 89.9) |  | 30.8 (28.6 a 33.0) | 59.2 (56.0 a 62.4) |  |
| **Growth and Development Control** |  |  |  |  |  |  |
| No | 39.2 (37.3 a 41.1) | 83.3 (81.1 a 85.4) | <0.001 | 39.8 (37.4 a 42.3) | 53.1 (50.2 a 56.0) | 0.456 |
| Yes | 60.8 (58.9 a 62.7) | 87.6 (86.2 a 88.9) |  | 60.2 (57.7 a 62.6) | 54.4 (52.3 a 56.6) |  |

\*Weighted estimate for the complex survey sample.
\*\*p-value calculated using the Rao-Scott test.

In the analysis of inequality in the completion of the two doses of measles vaccination mediated by wealth quintile (Figure 1), it was identified that mothers without education (CI: -0.57), aged 15 to 17 years (CI: -0.38), residing in the jungle region (CI: -0.41) and highlands (CI: -0.26), as well as rural areas (CI: -0.73) and without health insurance (CI: -0.01) showed greater inequality (Table 2). Meanwhile, infants without growth and development monitoring exhibited higher inequality in completing the two doses of measles vaccination (CI: -0.09).

**Table 2.** Characteristics of mothers and children and inequality in compliance with the two doses of measles vaccination.

| Variables | Complete Vaccination<br>Doses (n=3935) |  | Inequality in compliance with<br>the two doses of measles<br>vaccination against measles |  |
| --- | --- | --- | --- | --- |
|  | %* (95%CI) | P-value | CI (95%CI) | P-value |
| Mother's Characteristics |  |  |  |  |
| Age Group |  |  |  |  |
| 15 to 17 years old | 27 (13.0 a 47.8) | <0.001 | -0.376 (-0.665 a -0.086) | 0.011 |
| 18 to 29 years old | 47.5 (44.7 a 50.4) |  | -0.116 (-0.16 a -0.071) | <0.001 |
| 30 to 49 years old | 55.5 (53.2 a 57.7) |  | 0.067 (0.043 a 0.091) | <0.001 |
| Educational Level |  |  |  |  |
| No Education | 54.6 (40.8 a 67.7) | 0.380 | -0.573 (-0.763 a -0.383) | <0.001 |
| Elementary | 52.1 (48.1 a 56.2) |  | -0.515 (-0.576 a -0.455) | <0.001 |
| High School | 50.8 (48.2 a 53.4) |  | -0.163 (-0.198 a -0.128) | <0.001 |
| University | 53.7 (50.6 a 56.7) |  | 0.331 (0.3 a 0.361) | <0.001 |
| Natural Region |  |  |  |  |
| Metropolitan Lima | 53.3 (49.0 a 57.5) | 0.257 | 0.299 (0.26 a 0.339) | <0.001 |
| Rest of the Coast | 53.5 (50.7 a 56.3) |  | -0.023 (-0.081 a 0.036) | 0.449 |
| Highlands | 51.1 (48.6 a 53.7) |  | -0.261 (-0.319 a -0.204) | <0.001 |
| Jungle | 48.7 (45.7 a 51.8) |  | -0.414 (-0.469 a -0.359) | <0.001 |
| Area of Residence |  |  |  |  |
| Urban | 53.1 (50.9 a 55.2) | 0.004 | 0.118 (0.101 a 0.134) | <0.001 |
| Rural | 47.8 (45.0 a 50.6) |  | -0.725 (-0.774 a -0.676) | <0.001 |
| Health Insurance |  |  |  |  |
| Yes | 52.4 (50.5 a 54.3) | 0.300 | 0.198 (0.091 a 0.305) | <0.001 |
| No | 48.6 (41.8 a 55.6) |  | -0.014 (-0.023 a -0.005) | 0.002 |
| Antenatal Care Visits |  |  |  |  |
| Less than Six | 44.3 (39.3 a 49.3) | <0.001 | -0.078 (-0.163 a 0.007) | 0.073 |
| Six or More | 55.7 (53.7 a 57.6) |  | 0.01 (-0.001 a 0.02) | 0.063 |
| Received Tetanus Vaccine |  |  |  |  |
| No | 46.1 (42.2 a 49.9) | <0.001 | -0.003 (-0.006 a 0.001) | 0.152 |
| Yes | 56.2 (54.1 a 58.3) |  | -0.003 (-0.006 a 0.001) | 0.152 |
| Institutional Labor |  |  |  |  |
| No | 34.3 (28.4 a 40.7) | <0.001 | 0.022 (0.016 a 0.027) | <0.001 |
| Yes | 53 (51.2 a 54.9) |  | 0.022 (0.016 a 0.027) | <0.001 |
| Children's Characteristics |  |  |  |  |
| Age (months) |  |  |  |  |
| 12 to 23 | 29.1 (26.0 a 32.4) | <0.001 | -0.061 (-0.133 a 0.011) | 0.098 |
| 24 to 35 | 59.8 (56.2 a 63.4) |  | -0.033 (-0.087 a 0.02) | 0.226 |
| 36 to 47 | 58.7 (55.3 a 62.0) |  | 0.012 (-0.034 a 0.059) | 0.600 |
| 48 to 59 | 57.7 (54.4 a 60.9) |  | 0.042 (-0.005 a 0.088) | 0.079 |
| Growth and Development Control |  |  |  |  |
| No | 52.1 (49.1 a 55.0) | 0.823 | -0.094 (-0.124 a -0.064) | <0.001 |
| Yes | 52.5 (50.3 a 54.7) |  | 0.132 (0.091 a 0.173) | <0.001 |

**CI:** Concentration Index
\*Weighted estimate for the complex survey sample.

**Figure 1.**
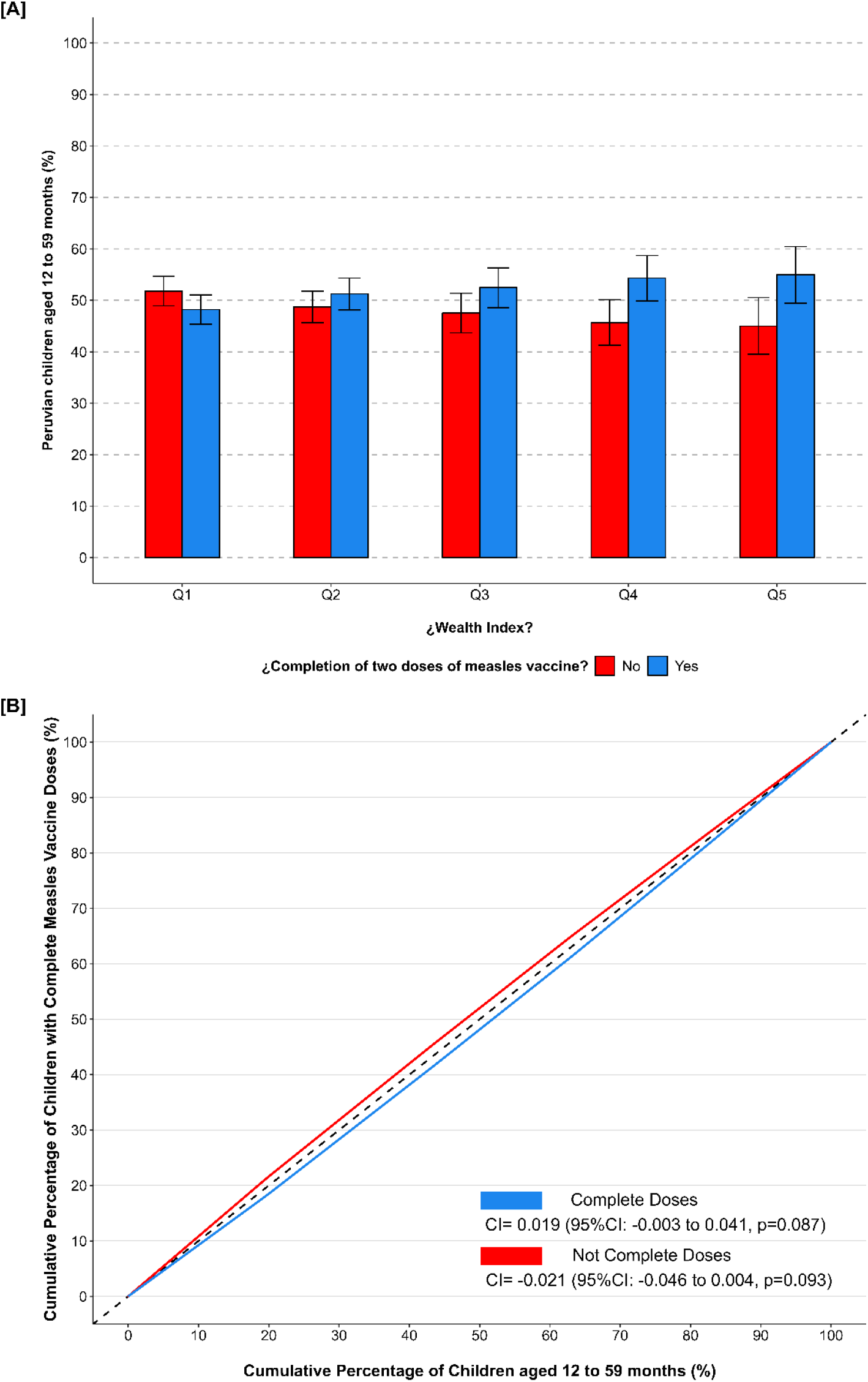
Inequalities in compliance with the two doses of measles vaccination in Peruvian children aged 12 to 59 months in 2023 **CI:** Concentration Index.

In assessing the prevalence of measles immunization, it was identified that children of women who had six or more prenatal care visits during pregnancy had a higher prevalence of 13.0% for vaccination with the first dose (aPR: 1.13, 95%CI: 1.07 – 1.19, p*<*0.001), as well as those children of mothers who had a higher prevalence of 7.8% for vaccination with the first dose during pregnancy (aPR: 1.08, 95%CI: 1.04 – 1.12, p*<*0.001). On the other hand, it was identified that children of mothers who had institutional childbirth had a higher prevalence of 10% (aPR: 1.10, 95%CI: 1.04 – 1.17, p=0.004) for measles immunization with the first dose. Similarly, it was found that the prevalence of immunization with the first dose increased as children got older, as well as among those who had growth and development check-ups (Table 3).

**Table 3.**
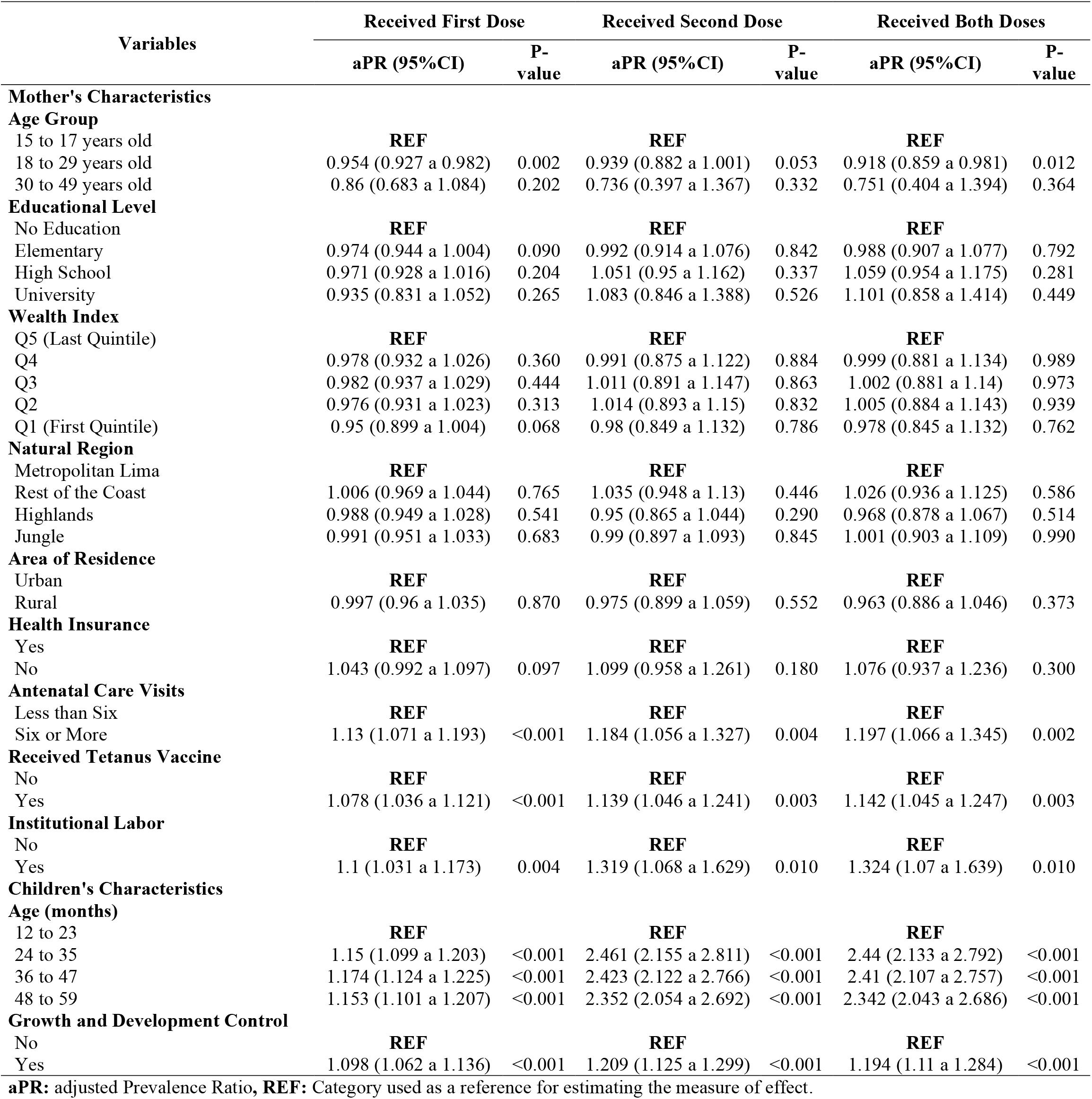
Association between mothers’ and children’s characteristics and compliance with two doses of measles vaccination.

In evaluating the prevalence of measles booster immunization, it was identified that children of women who had six or more prenatal care visits during pregnancy had a higher prevalence of 18.4% for vaccination (aPR: 1.18, 95%CI: 1.06 – 1.33, p=0.004), compared to children of women with fewer than six prenatal care visits. On the other hand, it was found that children of women who had institutional childbirth and received tetanus vaccination during pregnancy had a higher prevalence of 9.9% and 7.7%, respectively, for vaccination with the booster dose. Additionally, it was observed that the prevalence of immunization with the booster dose increased with children’s age, as well as among those who had growth and development check-ups (Table 3).

In assessing the prevalence of measles immunization with both doses, it was found that children of women who had six or more prenatal care visits during pregnancy had a higher prevalence of 19.7% for vaccination (PRa: 1.20, 95%CI: 1.07 – 1.35, p=0.002), compared to children of women with fewer than six prenatal care visits. On the other hand, it was identified that children of women who had institutional childbirth and received tetanus vaccination during pregnancy had a higher prevalence of 32.4% and 14.2%, respectively, for vaccination with both doses. Additionally, it was observed that the prevalence of immunization with both doses increased with children’s age, as well as among those who had growth and development checkups (Table 3).

In the assessment of inequality in measles immunization with one and/or two doses using the Erreygers Concentration Index (ECI), it was found that vaccination was concentrated in the lower wealth quintiles among children of women residing in the jungle region, without education, and who received tetanus vaccination during pregnancy. This pattern was also observed for children with growth and development monitoring. Conversely, the inequality in vaccination with both doses concentrated in the lower wealth quintiles was lower among children of women without education, with secondary education, and health insurance. Similarly, this was the case for children without growth and development monitoring (Table 4).

**Table 4.**
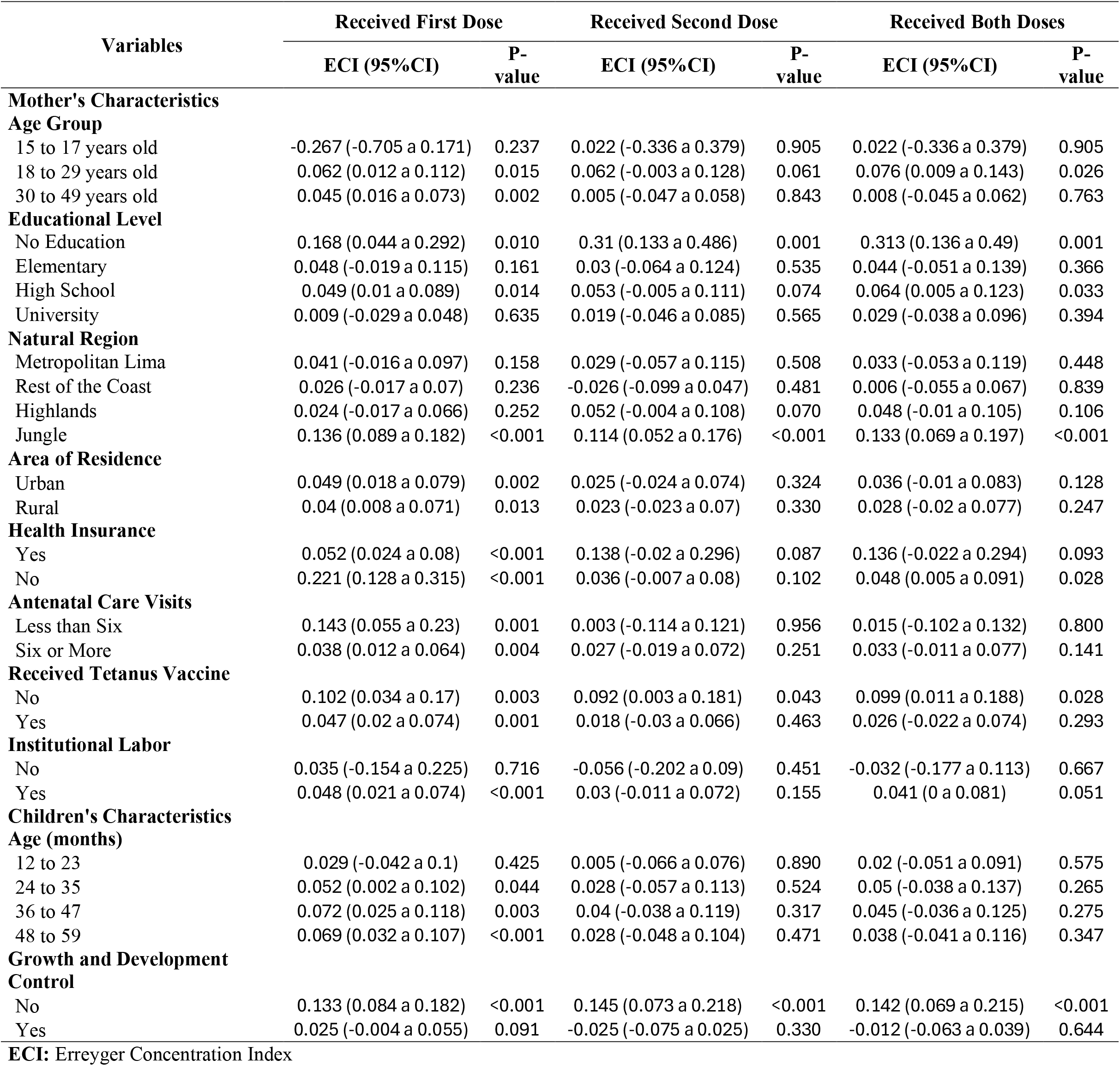
Inequality in compliance with the two doses of measles vaccination in children between 12 and 59 months of age.

In the assessment of measles immunization coverage across the 25 regions in Peru, it was identified that the departments of Apurímac, Ancash, Piura, and Tumbes had the highest immunization coverage. Conversely, the departments of Ucayali, Madre de Dios, Puno, and Amazonas had the lowest immunization coverage rates. Regarding inequality in immunization coverage with both doses of measles vaccine, it was found that the departments of Moquegua, Huancavelica, and Puno had higher inequality, whereas regions like Amazonas, Loreto, and Ucayali exhibited lower inequality (Figure 2).

**Figure 2.**
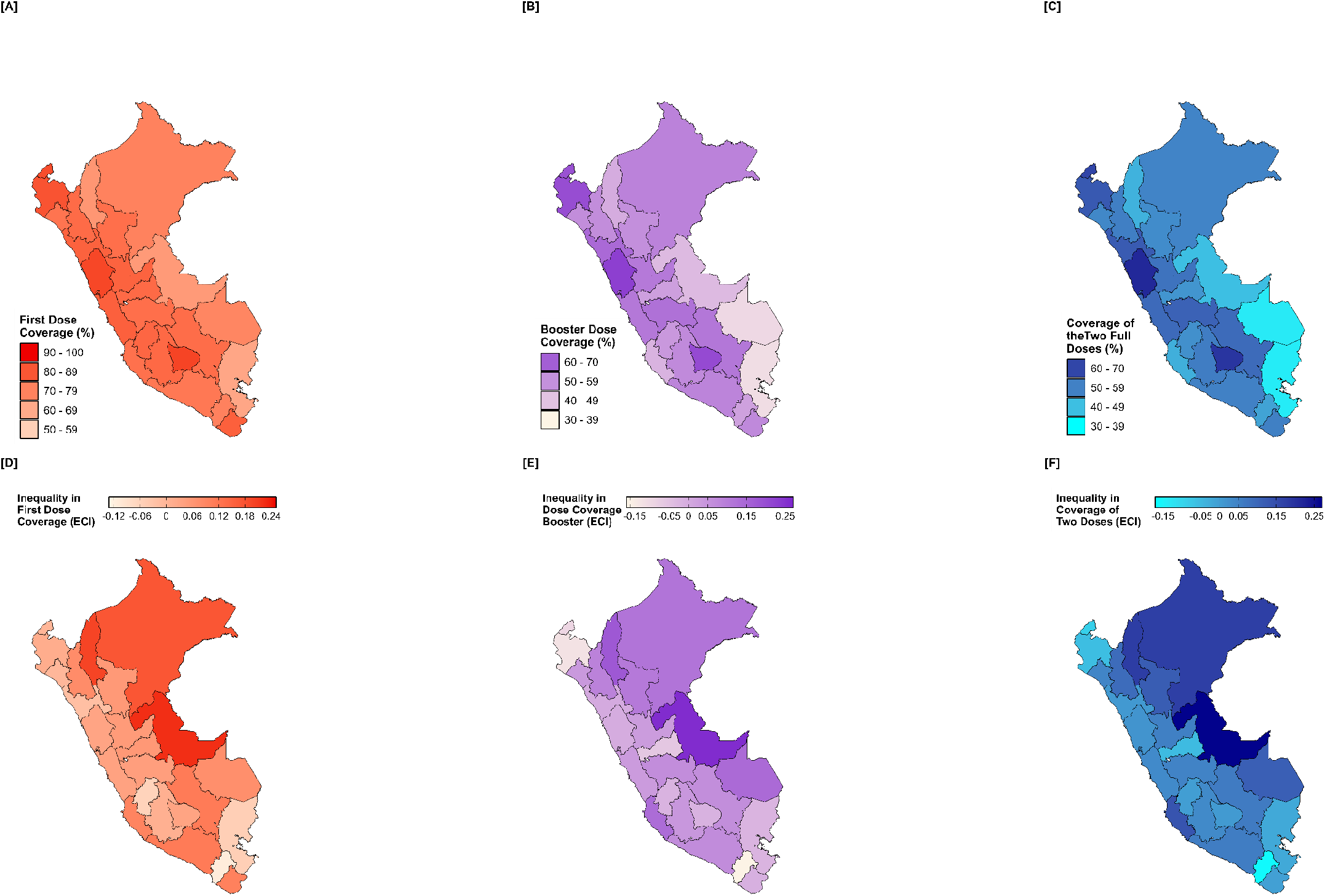
Geographical distribution of inequality in compliance with the two doses of measles vaccination in Peruvian children aged 12 to 59 months **ECI:** Erreygers Concentration Index

## DISCUSSION

This study examined inequalities in measles vaccination rates based on sociodemographic factors among Peruvian children aged 12 to 59 months. Our findings demonstrate inequality in both the first and second doses of vaccination, primarily mediated by sociodemographic factors such as maternal education level, age, and area of residence. Similarly, health-related factors such as the number of prenatal and growth and development check-ups for the child were associated with vaccination disparity.

Access to healthcare services in Peru is difficult, which worsens disparities in measles vaccination (18). In rural areas of the highlands and rainforests, the distance to vaccination centers forces the population to endure long journeys through strenuous walks or inadequate transportation means (19,20). This entails direct costs such as fares and consultation fees, as well as indirect costs like loss of income due to time spent traveling to vaccination sites (21). Therefore, this situation, coupled with poverty and low education levels, contributes to the abandonment of vaccination programs (22,23).

In contrast, urban areas enjoy privileged access to private healthcare services and have higher levels of health literacy (24,25). Moreover, due to the population density in these areas, they are the primary target of immunization campaigns (26,27). However, it is necessary to implement strategies such as verifying immunization in educational centers and disseminating information through social networks with clear messages in multiple languages to raise awareness among the population and improve vaccination coverage (28,29).

Mothers with higher formal education have children with less inequality in the coverage of both doses of measles vaccination. Mothers with higher education tend to better understand the importance of vaccination and have more resources available to vaccinate their children (30,31). Additionally, this group can more easily discern misinformation about vaccination risks in children (32). Therefore, education influences individual and collective decisions about vaccination, which can be improved through the implementation of educational strategies and policies that ensure equitable access to vaccines for the entire population (33).

Mothers with health insurance showed less inequality in completing measles vaccination doses. This is because the national immunization strategy for children under six years ensures free vaccine administration at all health facilities (34). Therefore, insured mothers can ensure completing their children’s vaccination schedule at different stages of their lives (35). However, some studies indicate that having health insurance may still lead to increased disparities in vaccination access, potentially because of differences in healthcare coverage and quality in countries like Peru (36-38).

Children enrolled in the Growth and Development Monitoring Program showed less inequality in completing the two doses of measles vaccination in Peru. This program offers child health assessments and emphasizes the importance of vaccination (39). Thus, even under unfavorable sociodemographic conditions, participation in this program can lead to higher immunization rates (40). However, conducting multiple activities during consultations and their limited duration may hinder discussions about the vaccination schedule (41). Strategies such as providing personalized information and sending reminders via phone calls or text messages have been shown to increase visits to health centers to complete the vaccination schedule (42,43).

In Peru, the average age of first pregnancy ranges from 21 to 23 years (44). Thus, a lack of experience and information among young pregnant women may reduce adherence to the immunization schedule (45,46). Additionally, young pregnant women are more exposed to biased information regarding the negative effects of vaccination on their children (47). An effective strategy has been training on infant health topics during prenatal check-ups (48). This setting is ideal due to the number of sessions and attentiveness provided by healthcare personnel (49), which enhances trust and satisfaction among pregnant women and increases their participation in postpartum vaccination programs (50).

Challenges such as insufficient supplies, healthcare system gaps, and poor monitoring limit childhood immunization (51,52). Therefore, prioritizing vaccination in areas with low coverage or outbreaks is crucial for preventing outbreaks (53). In low- and middle-income countries, knowledge of the vaccination schedule is often deficient due to a lack of records (54). Thus, informative sessions and a surveillance system can enhance immunization in healthcare centers (55). Therefore, assessing the effectiveness of these interventions in Peru is recommended to decrease the incidence of measles and other preventable diseases.

The study had some limitations because it was not possible to address mothers’ perceptions for not vaccinating their children or the difficulties they faced when seeking to complete the immunization schedule. Also, the regional level scope of the DHS prevents us from assessing coverage at the provincial and district levels to address the areas at highest risk of measles outbreaks in Peru.

In conclusion, inequalities in vaccination coverage with the first dose and booster dose against measles were greater among children aged 12 to 59 months of women living in rural areas or outside Lima Metropolitan area, lower education, and without health insurance. Additionally, vaccination rates were primarily concentrated among children whose mothers during pregnancy had six or more prenatal check-ups, received tetanus vaccines, and had institutional deliveries.

## Data Availability

The data obtained from the National Institute of Statistics and Informatics (or INEI, in Spanish) platform: https://proyectos.inei.gob.pe/microdatos/

## Acknowledgements

None.

